# Polygenic scores of subcortical brain volumes as possible modulators of treatment response in depression

**DOI:** 10.1101/2023.08.26.23294659

**Authors:** Vincenzo Oliva, Alfonso Martone, Giuseppe Fanelli, Katharina Domschke, Alessandra Minelli, Massimo Gennarelli, Paolo Martini, Marco Bortolomasi, Eduard Maron, Alessio Squassina, Claudia Pisanu, Siegfried Kasper, Joseph Zohar, Daniel Souery, Stuart Montgomery, Diego Albani, Gianluigi Forloni, Panagiotis Ferentinos, Dan Rujescu, Julien Mendlewicz, Diana De Ronchi, Bernhard T Baune, European College of Neuropsychopharmacology (ECNP) Pharmacogenomics & Transcriptomics Network, Alessandro Serretti, Chiara Fabbri

## Abstract

A significant proportion of patients with major depressive disorder (MDD) do not experience remission after one or more pharmacological treatments. Research has explored brain structural measures, particularly the hippocampus, as potential predictors of treatment response in MDD, as well as genetic factors.

This study investigated the association of polygenic scores (PGSs) for seven subcortical brain volumes (including the hippocampus, nucleus accumbens, amygdala, and caudate nucleus) with treatment non-response and non-remission in MDD.

Patients with MDD were recruited in the context of five clinical studies, including a total of 3,637 individuals. PGSs were estimated using a Bayesian framework and continuous shrinkage priors (PRS-CS-auto) after standard genotype quality control and imputation. Logistic regressions were performed between PGSs and non-response or non-remission in each sample, adjusting for age, sex, baseline symptom severity, recruitment sites, and population stratification. Results were meta-analysed across samples, using a random-effect model.

Caudate volume PGS was nominally associated with non-remission (OR=1.09, 95% CI=1.01–1.19, p=0.036). Leave-one-out sensitivity analyses suggested a possible association with the amygdala and thalamus PGSs. However, no association was significant after multiple testing correction.

These results, although preliminary, suggest a possible link between caudate volume PGS and lack of symptom remission. Methodological improvements in PGSs estimation and statistical power may enhance their predictive performance and provide a contribution to precision psychiatry.

## 1 Introduction

Major depressive disorder (MDD) is a prevalent psychiatric condition and one of the leading causes of disability worldwide, with a 61.1% increase in the number of disability-adjusted life years (DALYs) over the past two decades [1].

The first prescribed medication for depression may fail to produce symptom remission in up to 60% of patients, leading to therapy changes that follow a trial and error approach (e.g., switch to another drug, or augmentation with a different pharmacological agent) [2, 3]. Patients without adequate medication response have higher relapse rates [4]. Identifying the most suitable treatment for each patient early on, according to precision psychiatry [5, 6], may reduce the burden of the disease and the related costs to society [7].

Several socio-demographic and clinical factors have been recognised as important predictors of response and remission to psychopharmacological treatment in MDD, such as longer duration of depressive episodes, greater baseline severity, older age, and the presence of anxiety symptoms [8-10].

Common genetic variants were demonstrated to explain about 13% of the variability in remission [11], therefore polygenic scores (PGSs) represent a promising opportunity for investigating the genetic factors involved in treatment efficacy in MDD [12]. PGSs, alternatively called polygenic risk scores (PRSs) when they estimate the risk of a disease attributable to common variants [13], have also been used to understand disease pathogenesis [14], to identify the genetic overlap between different traits [15, 16], and they are promising potential predictors of treatment outcome [17]. Higher PRSs for MDD, schizophrenia (SCZ) and attention-deficit hyperactivity disorder were associated with a worse response to medications for depression [18, 19]. Interestingly, a low PRS for SCZ may decrease the benefits of augmentation with medications commonly employed as first-line treatments for SCZ [20]. PGSs for conscientiousness and neuroticism were other factors associated with response, while the PGSs of openness was inversely associated with remission and response [21]. PRSs for nonpsychiatric phenotypes, including PRSs for coronary artery disease, obesity, and cardioembolic stroke, were also inversely associated with response to treatment [22, 23].

The prediction of treatment response may be improved by considering PGSs of brain-related traits other than those expressing disease risk. Brain structural measures have been investigated as predictors of response to treatments in MDD. Alterations in the volumes of subcortical brain structures, particularly the hippocampus, have been among the most replicated findings [24]. Patients with MDD showed a volumetric reduction of the hippocampus vs healthy controls [25], and the hippocampal volume may distinguish treatment responders from non-responders [26]. The integrity of white matter tracts in the cortico-striatal-limbic systems was also implicated in predicting response to treatment [24]; these tracts connect the orbitofrontal cortex with subcortical structures, such as the nucleus accumbens, amygdala, caudate nucleus, globus pallidus, putamen, and thalamus [27]. The importance of subcortical structures in the modulation of response to medications for depression was also highlighted by functional studies showing that the activity of the amygdala may predict response to psychedelic drugs [28].

Genetic studies in family cohorts and twin studies have revealed varying levels of heritability for each subcortical structure, ranging from moderate to high. Heritability estimates based on single nucleotide polymorphisms (SNPs) were slightly lower, as expected, ranging from 17% to 47% for the thalamus and from 9% to 33% for the amygdala and brainstem, depending on the specific estimation method used [29-31]. However, the possible association between the polygenic component of brain subcortical structures and response to medications in MDD has not been investigated to the best of our knowledge. To contribute to fill this gap, we investigated the relationship between the PGSs for seven subcortical brain structure volumes (i.e., nucleus accumbens, amygdala, caudate nucleus, globus pallidus, putamen, thalamus, and hippocampus) and non-response and non-remission, across five clinical cohorts of patients with MDD. Our hypothesis was that the PGSs of these traits could be associated with medication treatment efficacy and could contribute to the future development of models able to aid the early identification of non-responders and non-remitters.

## 2 Material and methods

### 2.1 Target samples

#### 2.1.1 Brescia

This sample included 501 subjects with MDD (DSM-IV criteria) who were referred to the “Villa Santa Chiara” Psychiatric Hospital in Verona, Italy. The diagnosis was confirmed using the Structured Clinical Interview for DSM-IV Axis I Disorders (SCID-I). Participants who had other primary neuropsychiatric disorders, including intellectual disabilities, substance/alcohol abuse or dependence, dementias, or comorbid eating disorders, were excluded. The severity of symptoms was evaluated using the Montgomery-Asberg Depression Rating Scale (MADRS). Response was defined as ≥ 50% improvement in symptom severity during the current pharmacological treatment. Non-responders were patients who did not respond to at least one pharmacological treatment and were categorised as Stage I-III according to the Thase and Rush staging method [32]. Genome-wide genotyping was conducted using either the Infinium PsychArray-24 BeadChip or the Infinium Multi-Ethnic Genotyping Array (N = 215 and 286, respectively, denominated Brescia sample 1 and Brescia sample 2 in the supplementary materials). Further details are available elsewhere [33].

#### 2.1.2 European Group for the Study of Resistant Depression

A total of 1410 participants were recruited by the European Group for the Study of Resistant Depression (GSRD) as part of a multicentric cross-sectional study. Patients had a diagnosis of MDD (DSM-IV-TR criteria), according to the Mini International Neuropsychiatric Interview (MINI), and they were treated with a medication for depression for ≥4 weeks. The main exclusion criterion was another primary psychiatric disorder in the six months before enrolment. Depression severity was assessed using the MADRS at study entry and at the beginning of current episode (retrospectively, from anamnestic information and medical records). Treatment response was defined as a ≥50% reduction in the MADRS total score compared to the beginning of the current episode, after treatment with a medication for depression for ≥4 weeks. Non-responders were patients who did not respond to one or more pharmacological treatments during the current depressive episode. Remission was determined based on a current MADRS ≤10 after treatment with a medication for depression for ≥4 weeks. Genome-wide genotyping was performed with Infinium PsychArray-24 BeadChip. Additional information on this study is available elsewhere [34].

#### 2.1.3 Münster

This naturalistic study involved 621 individuals with MDD (DSM-IV criteria), confirmed using the SCID-I [35], recruited at the Department of Psychiatry, University of Münster, Germany. Patients with current alcohol/drug dependence or other primary neuropsychiatric disorders were excluded. Treatment response and remission were evaluated after six weeks of treatment, using the 21-item Hamilton Depression Rating Scale (HAMD_21_) (≥50% improvement from baseline and HAMD_21_ ≤7, respectively). Genome-wide genotyping was conducted using the Infinium PsychArray-24 BeadChip.

#### 2.1.4 Sequenced treatment alternatives to relieve depression

The Sequenced treatment alternatives to relieve depression (STAR*D) study evaluated the effectiveness and tolerability of different medications in treating moderate-severe MDD in primary care or psychiatric outpatient clinics. Patients with any other primary psychiatric diagnosis were excluded. The study initially involved treatment with citalopram for 12 weeks, and only this phase of the study was considered for the present analyses. Symptom severity was assessed using the Quick Inventory of Depressive Symptomatology Clinician-rated scale (QIDS-C_16_). Response was defined as a ≥ 50% decrease in symptom severity after 12 weeks of treatment with citalopram, and remission as QIDS-C_16_ ≤5 at week 12. A total of 1,948 participants were genome-wide genotyped using the Affymetrix GeneChip Human Mapping 500 K Array Set or Affymetrix Genome-Wide Human SNP Array 5.0. Further details of the study are available elsewhere [36].

#### 2.1.5 Tartu

This sample included 83 outpatients with MDD (DSM-IV criteria) recruited at the Psychiatric Clinic of the University Hospital of Tartu, Estonia. The diagnosis was verified using medical records and MINI 5.0.0. Patients with current suicide risk or another primary neuropsychiatric disorder were excluded. Treatment response was defined as a ≥50% decrease in MADRS scores from baseline, while remission as MADRS ≤10 at week six. The Illumina 370CNV array was used for genome-wide genotyping. Additional details are available elsewhere [37].

### 2.2 Quality control of genotypes in the target datasets

Each of the five target samples underwent quality control (QC) and population principal component analysis (PCA) through the Ricopili pipeline [38]. SNPs were filtered retaining those with call rate ≥0.95, missing difference between cases-controls ≤0.02, minor allele frequency (MAF)≥0.01, and Hardy-Weinberg equilibrium p-value ≥1e-6. Individuals were retained if they had an autosomal heterozygosity deviation within ±0.2, call rate ≥0.98, and no genetic/phenotypic sex mismatch.

To assess between-subjects relatedness and population stratification, linkage disequilibrium-pruned data (R2<0.2) were used to identify all pairs of individuals with identity-by-descent proportion >0.2, and one individual from each pair was removed. Population stratification was determined using PCA (Eigenstrat); population outliers were removed according to the mean ±6 standard deviations of the first 20 principal components (PCs). Individuals of European ancestry were retained based on self-report/anamnestic information and inspection of PCA plots [38].

Genotype imputation was carried out on the Michigan Imputation Server [39] using Minimac4 and the Haplotype Reference Consortium (HRC) r1.1 2016 (GRCh37/hg19).

Post-imputation QC was performed by filtering out variants having a poor imputation quality score (R^2^<0.3) and MAF<0.05.

### 2.3 Statistical analyses

Summary statistics of the largest available GWASs on subcortical brain structure volumes were used as base datasets [30, 31]. We computed PGSs using PRS-CS-auto, a Bayesian method that places continuous shrinkage priors on SNP effect sizes and obviates the need to select any *a priori* GWAS P-threshold for SNP inclusion [40].

Logistic regressions between each scaled PGS (mean=0, SD=1) and each clinical outcome (non-response and non-remission) were conducted using R v4.0.2, adjusting for age, sex, baseline symptom severity (for non-remission), relevant population principal components, and recruitment sites, similarly to a previous study [18]. The variance in the outcomes explained by each PGS was estimated as the difference between the Nagelkerke’s pseudo R^2^ of the full models and those including the covariates only, in each cohort separately [15].

The results obtained in each sample were meta-analysed using the R metafor package [41], within a random-effects model, using the restricted maximum-likelihood estimator [42]. Analyses of heterogeneity were performed using the Cochran’s Q test [43], and I^2^ statistic (0% indicates no observed heterogeneity, and higher values indicate increasing heterogeneity, with 25%, 50%, and 75% defining thresholds for low, moderate, and high) [44]. Leave-one-out sensitivity analyses were performed to investigate the influence of each individual study on the results, by omitting one study at a time [45].

The Bonferroni correction was applied considering the seven base phenotypes analysed (α=0.05/7=0.007).

We estimated statistical power using the AVENGEME R package [46]. Assuming a covariance of 50% between the base and target phenotypes, all the analysed PGSs showed adequate power (between 90% and 100%) for both target phenotypes. The power decreased to a range of 24%–48% when the covariance was set to 25%. Details on the power analysis are provided in the Supplementary materials.

## 3 Results

After QC, a total of 3,637 patients with MDD were included in the analyses (Brescia n=453; GSRD n=1149; Munster n=557; STAR*D n=1400; Tartu n=78). The description of the clinical-demographic characteristics of the target samples is provided elsewhere [18].

In the meta-analyses, no association survived after the Bonferroni correction (Table 1). The top result was found for the caudate nucleus PGS, which was nominally associated with non-remission (OR=1.09, 95% CI=1.01–1.19, p=0.036, range pseudo-R^2^=0.01-0.6%), with no evidence of heterogeneity (Table 1, a forest plot is depicted in Figure 1). Leave-one-out sensitivity analyses conducted for this association identified a significant influence of GSRD and STAR*D samples on the overall result; removing each one of these samples from the meta-analysis showed indeed an impact on the results (p=0.230 and p=0.351, respectively). For the other PGSs of interest, we also performed leave-one-out sensitivity analyses. After removing the Münster sample, there was a nominally significant association between the PGS of amygdala and both non-response (OR=1.09, 95% CI=1.01-1.19, p=0.048, range pseudo-R2=0.001-1.6%) and non-remission (OR=1.10, 95% CI=1.01-1.22, p=0.041, range pseudo-R2=0.11-1.4%). The association between thalamus PGS and non-remission became nominally significant when removing the STARD*D sample from the meta-analysis (OR=0.85, 95% CI=0.76-0.96, p=0.009, range pseudo-R2=0.006-1.4%). The results of the other leave-one-out sensitivity analyses and of the regression analyses in each sample are provided in the Supplementary materials.

**Table 1.**
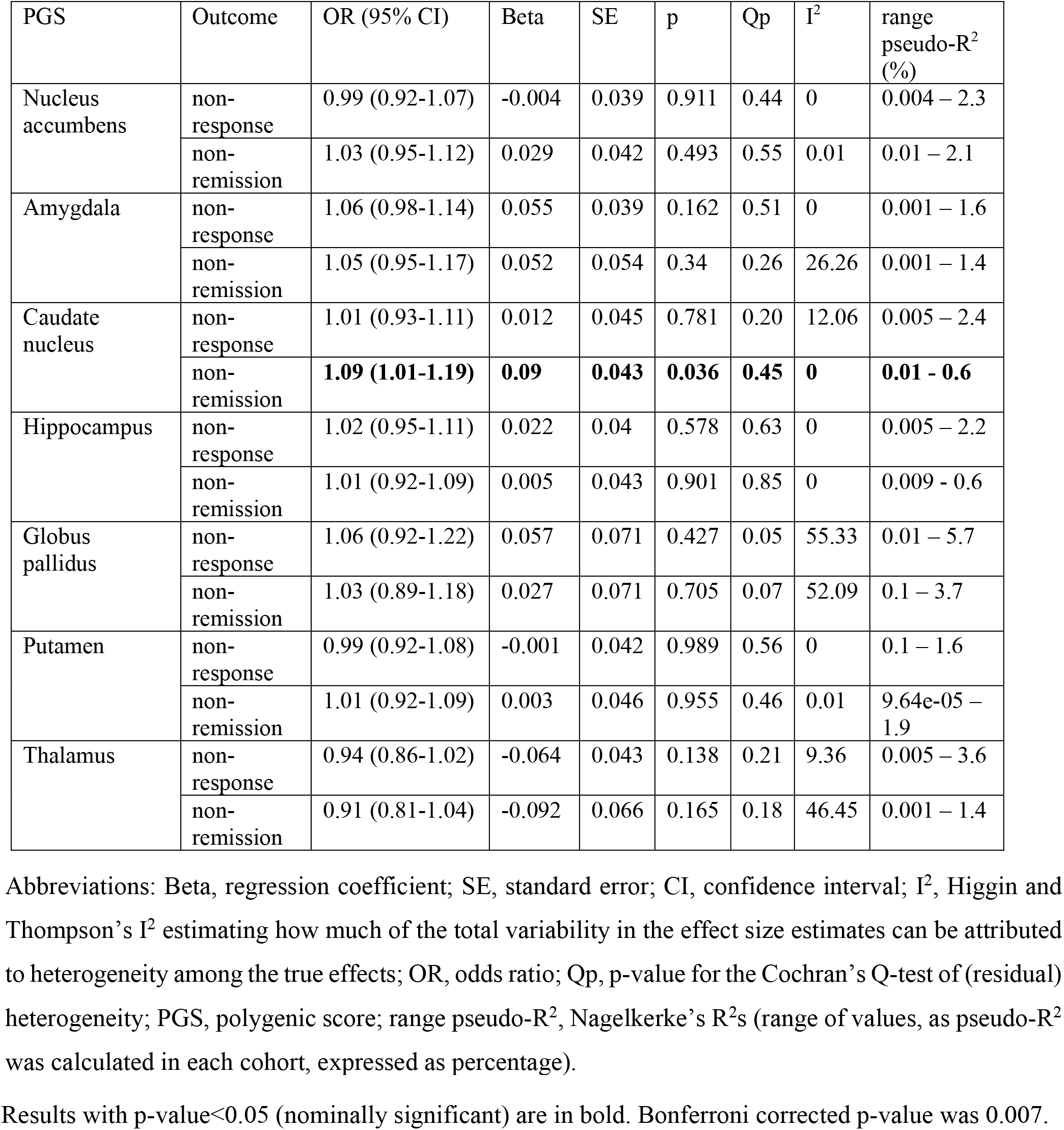
Results of the meta-analyses.

| PGS | Outcome | OR (95% CI) | Beta | SE | p | Qp | I<sup>2</sup> | range pseudo-R<sup>2</sup> (%) |
| --- | --- | --- | --- | --- | --- | --- | --- | --- |
| Nucleus accumbens | non-response | 0.99 (0.92-1.07) | -0.004 | 0.039 | 0.911 | 0.44 | 0 | 0.004 – 2.3 |
|  | non-remission | 1.03 (0.95-1.12) | 0.029 | 0.042 | 0.493 | 0.55 | 0.01 | 0.01 – 2.1 |
| Amygdala | non-response | 1.06 (0.98-1.14) | 0.055 | 0.039 | 0.162 | 0.51 | 0 | 0.001 – 1.6 |
|  | non-remission | 1.05 (0.95-1.17) | 0.052 | 0.054 | 0.34 | 0.26 | 26.26 | 0.001 – 1.4 |
| Caudate nucleus | non-response | 1.01 (0.93-1.11) | 0.012 | 0.045 | 0.781 | 0.20 | 12.06 | 0.005 – 2.4 |
|  | non-remission | **1.09 (1.01-1.19)** | **0.09** | **0.043** | **0.036** | **0.45** | **0** | **0.01 - 0.6** |
| Hippocampus | non-response | 1.02 (0.95-1.11) | 0.022 | 0.04 | 0.578 | 0.63 | 0 | 0.005 – 2.2 |
|  | non-remission | 1.01 (0.92-1.09) | 0.005 | 0.043 | 0.901 | 0.85 | 0 | 0.009 - 0.6 |
| Globus pallidus | non-response | 1.06 (0.92-1.22) | 0.057 | 0.071 | 0.427 | 0.05 | 55.33 | 0.01 – 5.7 |
|  | non-remission | 1.03 (0.89-1.18) | 0.027 | 0.071 | 0.705 | 0.07 | 52.09 | 0.1 – 3.7 |
| Putamen | non-response | 0.99 (0.92-1.08) | -0.001 | 0.042 | 0.989 | 0.56 | 0 | 0.1 – 1.6 |
|  | non-remission | 1.01 (0.92-1.09) | 0.003 | 0.046 | 0.955 | 0.46 | 0.01 | 9.64e-05 – 1.9 |
| Thalamus | non-response | 0.94 (0.86-1.02) | -0.064 | 0.043 | 0.138 | 0.21 | 9.36 | 0.005 – 3.6 |
|  | non-remission | 0.91 (0.81-1.04) | -0.092 | 0.066 | 0.165 | 0.18 | 46.45 | 0.001 – 1.4 |

Abbreviations: Beta, regression coefficient; SE, standard error; CI, confidence interval; I<sup>2</sup>, Higgin and Thompson's I<sup>2</sup> estimating how much of the total variability in the effect size estimates can be attributed to heterogeneity among the true effects; OR, odds ratio; Qp, p-value for the Cochran's Q-test of (residual) heterogeneity; PGS, polygenic score; range pseudo-R<sup>2</sup>, Nagelkerke's R<sup>2</sup>s (range of values, as pseudo-R<sup>2</sup> was calculated in each cohort, expressed as percentage).
Results with p-value<0.05 (nominally significant) are in bold. Bonferroni corrected p-value was 0.007.

**Figure 1.**
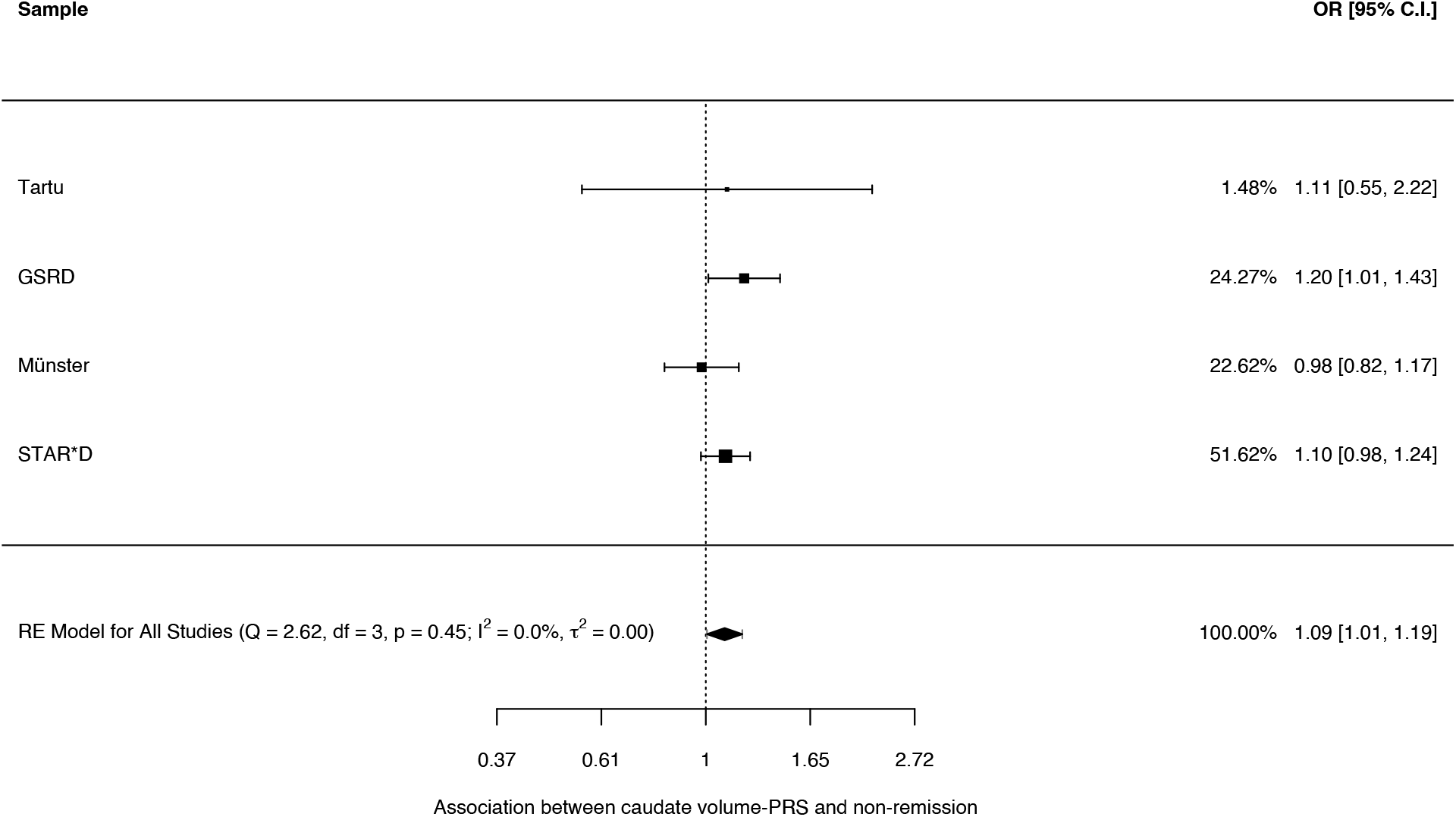
Forest plot showing the association between the polygenic score for major depressive disorder and non-remission.

## 4 Discussion

In the present meta-analysis, we investigated whether the PGSs for seven brain subcortical volumes were associated with non-response or non-remission in a sample of 3,637 patients with MDD. A nominal association was found between caudate nucleus PGS and non-remission. Leave-one-out analyses showed nominal associations between a higher PGS for amygdala volume and both non-response and non-remission, and between a lower PGS for thalamus volume and non-remission. However, no association survived after Bonferroni correction. Previous evidence of associations between hippocampal volume, MDD and response to pharmacological treatment [25, 26] were not corroborated by our study, as no association between hippocampal volume PGS and response/remission was found.

Though it did not survive a strict multiple-testing correction, a higher PGS for caudate nucleus volume may be associated with non-remission, an effect that was independent of age, sex, and baseline symptom severity. The caudate nucleus is important for executive function, which includes the regulation of affective states, hence the interest in this structure in the study of MDD [47]. A previous meta-analysis of neuroimaging studies reported significant volume reductions in the caudate nucleus in patients with depression compared to healthy controls [48], which were attributed to decreases in both grey matter volume [49] and neuronal density [50]. Greater caudate reduction was associated with more severe depression [51]. Preliminary evidence from small samples indicated that caudate nuclei do not differ between non-responders and responders to pharmacological treatment, although possible sex effects were suggested [52]. However, fronto-striatal atrophy, characterized by a reduction in caudate volume, was observed in patients with treatment-resistant depression when compared to patients with depression in remission [53]. Additionally, reduced bilateral caudate volume was observed in patients with MDD and elevated anhedonic symptoms [54], which are also associated with treatment non-response [24]. The caudate nucleus has a key role in the reward system [55], and anhedonia is believed to result from the dysfunction of reward and motivational dopaminergic neural circuits [56]. A PET study investigated dopaminergic receptor availability in the striatum, including the caudate nucleus, in individuals with MDD. Patients, particularly non-remitters, demonstrated higher dopaminergic receptor availability compared to healthy controls, suggesting a significant role of dopaminergic dysfunction in response to first-line treatments for depression [57]. Although our findings are apparently not in line with some previous evidence from brain imaging studies, a recent work showed positive genetic correlation between MDD and caudate volume [58], therefore genetic factors associated with higher volume of this structure may overlap with both MDD and lack of remission to treatment. Environmental factors interact with genetic variables in determining brain volumes, therefore future studies should also consider the modulating effects of the environment on genetic factors. Taken collectively, these findings suggest the potential relevance of augmentation therapy employing drugs that target the dopaminergic system for MDD [59]. However, we underline that our result did not survive multiple-testing correction, and it was influenced by the two largest samples in the meta-analysis (i.e., GSRD and STAR*D). Independent studies in samples of sufficient size and adequate power are important to verify our results.

The associations between the PGS for amygdala volume and both response and remission became nominally significant after removing the Münster sample from the main analysis. This sample is the only one among those included in the meta-analysis that used the HAMD_21_ for the assessment of response and remission. The validity of this scale in assessing treatment efficacy has been questioned, due to its low sensibility to change; its widespread use in clinical trials was suggested to have hampered the detection of significant placebo-medication differences [60]. Therefore, it is possible that the use of this scale in the Münster sample may have influenced the comparability with the other cohorts and affected our results. The amygdala has long been an interesting region in neurobiological studies of depressive-anxious disorders, owing to its relevant role in emotional processing and regulation [61]. A previous neuroimaging meta-analysis found no significant difference in amygdala volume between people with depression and healthy controls; however, the inclusion of only patients taking medications in the case group may have affected the results [62]. Neuroimaging functional studies showed a link between non-remission and lower fractional anisotropy in white matter pathways to the amygdala [26] and decreased amygdala function [63], whereas better response was associated with increased pre-treatment amygdala activity during the processing of emotional stimuli [64].

A nominally significant association between a lower PGS for thalamus volume and non-remission was observed only upon the exclusion of the STAR*D sample from the meta-analysis. As stated earlier, STAR*D was the largest sample in this analysis, and this could affect the results. Furthermore, the STAR*D sample differs from others in several ways. Firstly, it was the only study to use the QIDS_16_ for assessing efficacy outcomes. Secondly, the participants largely come from primary care settings, including individuals with long duration of illness, and significant medical and psychiatric comorbidities. Lastly, detailed assessments of medication adherence were not conducted, potentially leading to misclassification of outcomes when non-adherence to treatment contributes to a worse outcome [65]. The thalamus is a complex sensory information processing center that regulates emotion, memory, and arousal, and is composed of multiple anatomical subdivisions [66]. There is no consistent evidence supporting structural changes in the thalamus in MDD. An earlier meta-analysis of structural studies, including both treated and untreated patients with MDD with varying numbers of episodes, showed a significant reduction in gray matter volume in the thalamus in MDD patients [67]. However, a subsequent meta-analysis that focused only on untreated MDD patients revealed an increase in gray matter volume in the thalamus [68]. These findings suggest that medication use, number of depressive episodes, and/or underlying genetic factors may have an impact on the gray matter structure of the thalamus.

The strengths and limitations of this study should be considered. We used a standardised genetic quality control procedure on the target datasets in line with current standards, we applied a strict correction for multiple testing, performed a random-effect meta-analysis of our results across the five target samples, and included socio-demographic and clinical variables as potential confounders in the regression models. Potential limitations were the heterogeneity in the time points used for the assessment of efficacy outcomes in the target samples (i.e., six weeks, at least four weeks, or 12 weeks, depending on the cohort) and different medications across the samples (i.e., naturalistic treatment or citalopram in the case of STAR*D). Furthermore, distinguishing between non-response and non-remission outcomes may yield misleading results due to potential overlap with treatment-resistance. Future studies should aim to obtain a better harmonization of non-response, non-remission, and treatment-resistance phenotype definitions [69]. Additionally, the disparity in statistical power across different covariance estimates suggests a probable lack of sufficient power to detect significant results, emphasising the necessity of conducting studies with larger sample sizes to confirm and validate our findings. However, the proportion of phenotypic variance explained by PGSs was limited, and no association survived after multiple testing correction, suggesting that the PGSs of the examined brain subcortical volumes are unlikely to play a substantial role in response/remission.

In conclusion, our study found a nominally significant association between non-remission and the PGS for caudate nucleus volume. However, the examined PGSs were not able to predict treatment response or remission after multiple-testing correction. Future research could advance our knowledge in this field through methodological improvements in PGSs estimation, statistical power, and sample phenotyping.

## Statement of Ethics

The authors assert that all procedures contributing to this work comply with the ethical standards of the relevant national and institutional committees on human experimentation and with the Helsinki Declaration of 1975, as revised in 2008. All participants were included after obtaining their written informed consent. Brescia study protocol was approved by the Ethics Committee of the province of Verona, Italy; the European Group for the Study of Resistant Depression (GRSD) study protocol was approved by the Ethics Committee of the coordinating center at Hôpital Erasme, Cliniques universitaires de Bruxelles (Université Libre de Bruxelles), Belgium, and the local ethical committees of all the other nine participating centres; the Münster study protocol was approved by the ethical board of the University of Münster, Germany; the Tartu study protocol was approved by the Human Studies Ethics Committee of the University of Tartu and State Agency of Medicines; the Sequenced Treatment Alternatives to Relieve Depression (STAR*D) study protocol received ethics approval from 14 participating institutional review boards, a National Coordinating Center, a Data Coordinating Center, and the Data Safety and Monitoring Board at the National Institute of Mental Health (NIMH), Bethesda, Maryland, US.

## Authors Statement Contributors

Vincenzo Oliva contributed to the conceptualisation of the study, performed the analyses, interpreted the results and wrote the first draft of the manuscript. Alfonso Martone contributed to data analyses. Giuseppe Fanelli conceptualised the study, performed quality control and imputation of individual genotype data in each target sample, and reviewed the first draft of the manuscript. Alessandro Serretti and Chiara Fabbri conceptualised the study, helped with the interpretation of the results, reviewed the first draft of the manuscript. Chiara Fabbri created the polygenic scores and supervised the whole process leading to the final publication. The other authors contributed to data collection, data preparation and/or the improvement of the final version of the paper. All named authors meet the International Committee of Medical Journal Editors (ICMJE) criteria for authorship for this manuscript, take responsibility for the integrity of the work as a whole, and have given final approval for the version to be published.

## Acknowledgements

The European College of Neuropsychopharmacology (ECNP) Pharmacogenomics & Transcriptomics Network contributed to this manuscript, by sharing sample data, as well as providing comments and critical review to the manuscript.

We thank the US National Institute of Mental Health (NIMH) for providing access to data on the STAR∗D sample. We also thank the authors of previous publications in this dataset, and foremost, we thank the patients and their families who agreed to be enrolled in the study. Data were obtained from the limited access dataset distributed from the NIH-supported STAR∗D (NIMH Contract No. N01MH90003, ClinicalTrials.gov identifier is NCT00021528).

We thank the European Group for the Study of Resistant Depression (GRSD) for providing access to the dataset they collected, as well as the researchers of the consortia that provided the GWAS summary statistics used in our analyses and the participants of the cohorts to which they refer.

## Data Availability

Access to the Sequenced Treatment Alternatives to Relieve Depression (STAR∗D) data is granted to Principal Investigators after approval of a research proposal to the National Institute of Mental Health (NIMH) via the NIMH Repository & Genomics Resource (NRGR) (https://www.nimhgenetics.org).

## Declaration of Generative AI and AI-assisted technologies in the writing process

None.

## ECNP Network Transcriptomics & Pharmacogenomics, contributors

Marie-Claude Potier (Pitié-Salpêtrière Hospital, Paris, France)

Roos van Westrhenen (Outpatient Clinic Pharmacogenetics, Parnassia Psychiatric Institute/PsyQ, Amsterdam, Netherlands; Institute of Psychiatry, Psychology & Neuroscience (IoPPN), King’s College London, United Kingdom; School for Mental Health and Neuroscience FHML Maastricht, Netherlands)

Filip Rybakowski (Poznan University of Medical Science, Poznan, Poland)

Divya Mehta (Queensland University of Technology, Brisbane, Australia)

Mara Dierssen (Centre for Genomic Regulation, Barcelona, Spain)

Joost G.E. Janzing (Radboud University Nijmegen, Nijmegen, The Netherlands)

Pietro Lio’ (University of Cambridge, Cambridge, United Kingdom)

## References

[1] T. Vos, S. S. Lim, C. Abbafati, K. M. Abbas, M. Abbasi, M. Abbasifard, M. Abbasi-Kangevari, H. Abbastabar, F. Abd-Allah, and A. Abdelalim, “Global burden of 369 diseases and injuries in 204 countries and territories, 1990–2019: a systematic analysis for the Global Burden of Disease Study 2019,” The Lancet, vol. 396, no. 10258, pp. 1204–1222, 2020.

[2] V. De Carlo, R. Calati, and A. Serretti, “Socio-demographic and clinical predictors of non-response/non-remission in treatment resistant depressed patients: A systematic review,” Psychiatry Research, vol. 240, pp. 421–430, 2016/06/30/ 2016, doi: 10.1016/j.psychres.2016.04.034.

[3] R. van Westrhenen and M. Ingelman-Sundberg, “Editorial: From Trial and Error to Individualised Pharmacogenomics-Based Pharmacotherapy in Psychiatry,” (in English), Frontiers in Pharmacology, Editorial vol. 12, 2021-September-22 2021, doi: 10.3389/fphar.2021.725565.

[4] A. J. Rush, M. H. Trivedi, S. R. Wisniewski, A. A. Nierenberg, J. W. Stewart, D. Warden, G. Niederehe, M. E. Thase, P. W. Lavori, B. D. Lebowitz, P. J. McGrath, J. F. Rosenbaum, H. A. Sackeim, D. J. Kupfer, J. Luther, and M. Fava, “Acute and longer-term outcomes in depressed outpatients requiring one or several treatment steps: a STAR*D report,” (in eng), Am J Psychiatry, vol. 163, no. 11, pp. 1905–17, Nov 2006, doi: 10.1176/ajp.2006.163.11.1905.

[5] P. Fusar-Poli, M. Manchia, N. Koutsouleris, D. Leslie, C. Woopen, M. E. Calkins, M. Dunn, C. L. Tourneau, M. Mannikko, T. Mollema, D. Oliver, M. Rietschel, E. Z. Reininghaus, A. Squassina, L. Valmaggia, L. V. Kessing, E. Vieta, C. U. Correll, C. Arango, and O. A. Andreassen, “Ethical considerations for precision psychiatry: A roadmap for research and clinical practice,” (in eng), Eur Neuropsychopharmacol, vol. 63, pp. 17–34, Oct 2022, doi: 10.1016/j.euroneuro.2022.08.001.

[6] R. Zanardi, D. Prestifilippo, C. Fabbri, C. Colombo, E. Maron, and A. Serretti, “Precision psychiatry in clinical practice,” (in eng), Int J Psychiatry Clin Pract, vol. 25, no. 1, pp. 19–27, Mar 2021, doi: 10.1080/13651501.2020.1809680.

[7] A. Serretti, “Psychopharmacology: past, present and future,” (in eng), Int Clin Psychopharmacol, vol. 37, no. 3, pp. 82–83, May 1 2022, doi: 10.1097/yic.0000000000000402.

[8] A. Kautzky, M. Dold, L. Bartova, M. Spies, T. Vanicek, D. Souery, S. Montgomery, J. Mendlewicz, J. Zohar, and C. Fabbri, “Refining prediction in treatment-resistant depression: results of machine learning analyses in the TRD III sample,” The Journal of clinical psychiatry, vol. 79, no. 1, p. 14989, 2017.

[9] P. Olgiati, G. Fanelli, and A. Serretti, “Obsessive-compulsive symptoms in major depressive disorder correlate with clinical severity and mixed features,” (in eng), Int Clin Psychopharmacol, vol. 37, no. 4, pp. 166–172, Jul 1 2022, doi: 10.1097/yic.0000000000000396.

[10] K. Domschke, J. Deckert, V. Arolt, and B. T. Baune, “Anxious versus non-anxious depression: difference in treatment outcome,” (in eng), J Psychopharmacol, vol. 24, no. 4, pp. 621–2, Apr 2010, doi: 10.1177/0269881108097723.

[11] O. Pain, K. Hodgson, V. Trubetskoy, S. Ripke, V. S. Marshe, M. J. Adams, E. M. Byrne, A. I. Campos, T. Carrillo-Roa, and A. Cattaneo, “Identifying the common genetic basis of antidepressant response,” Biological psychiatry global open science, vol. 2, no. 2, pp. 115–126, 2022.

[12] S. W. Choi and P. F. O’Reilly, “PRSice-2: Polygenic Risk Score software for biobank-scale data,” Gigascience, vol. 8, no. 7, p. giz082, 2019.

[13] J. E. Craig, X. Han, A. Qassim, M. Hassall, J. N. Cooke Bailey, T. G. Kinzy, A. P. Khawaja, J. An, H. Marshall, P. Gharahkhani, R. P. Igo, S. L. Graham, P. R. Healey, J.-S. Ong, T. Zhou, O. Siggs, M. H. Law, E. Souzeau, B. Ridge, P. G. Hysi, K. P. Burdon, R. A. Mills, J. Landers, J. B. Ruddle, A. Agar, A. Galanopoulos, A. J. R. White, C. E. Willoughby, N. H. Andrew, S. Best, A. L. Vincent, I. Goldberg, G. Radford-Smith, N. G. Martin, G. W. Montgomery, V. Vitart, R. Hoehn, R. Wojciechowski, J. B. Jonas, T. Aung, L. R. Pasquale, A. J. Cree, S. Sivaprasad, N. A. Vallabh, A. C. Viswanathan, F. Pasutto, J. L. Haines, C. C. W. Klaver, C. M. van Duijn, R. J. Casson, P. J. Foster, P. T. Khaw, C. J. Hammond, D. A. Mackey, P. Mitchell, A. J. Lotery, J. L. Wiggs, A. W. Hewitt, S. MacGregor, N. consortium, U. K. B. Eye, and C. Vision, “Multitrait analysis of glaucoma identifies new risk loci and enables polygenic prediction of disease susceptibility and progression,” Nature Genetics, vol. 52, no. 2, pp. 160–166, 2020/02/01 2020, doi: 10.1038/s41588-019-0556-y.

[14] C. Fabbri, S. Montgomery, C. M. Lewis, and A. Serretti, “Genetics and major depressive disorder: clinical implications for disease risk, prognosis and treatment,” International Clinical Psychopharmacology, vol. 35, no. 5, 2020. [Online]. Available: https://journals.lww.com/intclinpsychopharm/Fulltext/2020/09000/Genetics_and_major_depressive_disorder clinical.1.aspx.

[15] V. Oliva, G. Fanelli, S. Kasper, J. Zohar, D. Souery, S. Montgomery, D. Albani, G. Forloni, P. Ferentinos, D. Rujescu, J. Mendlewicz, D. De Ronchi, C. Fabbri, and A. Serretti, “Melancholic features and typical neurovegetative symptoms of major depressive disorder show specific polygenic patterns,” Journal of Affective Disorders, vol. 320, pp. 534–543, 2023/01/01/ 2023, doi: 10.1016/j.jad.2022.10.003.

[16] G. Fanelli, M. Sokolowski, D. Wasserman, E. C. o. N. N. o. S. Research, Prevention, S. Kasper, J. Zohar, D. Souery, S. Montgomery, D. Albani, and G. Forloni, “Polygenic risk scores for neuropsychiatric, inflammatory, and cardio-metabolic traits highlight possible genetic overlap with suicide attempt and treatment-emergent suicidal ideation,” American Journal of Medical Genetics Part B: Neuropsychiatric Genetics, vol. 189, no. 3-4, pp. 74–85, 2022.

[17] P. Natarajan, R. Young, N. O. Stitziel, S. Padmanabhan, U. Baber, R. Mehran, S. Sartori, V. Fuster, D. F. Reilly, and A. Butterworth, “Polygenic risk score identifies subgroup with higher burden of atherosclerosis and greater relative benefit from statin therapy in the primary prevention setting,” Circulation, vol. 135, no. 22, pp. 2091–2101, 2017.

[18] G. Fanelli, K. Domschke, A. Minelli, M. Gennarelli, P. Martini, M. Bortolomasi, E. Maron, A. Squassina, S. Kasper, and J. Zohar, “A meta-analysis of polygenic risk scores for mood disorders, neuroticism, and schizophrenia in antidepressant response,” European Neuropsychopharmacology, vol. 55, pp. 86–95, 2022.

[19] C. Fabbri, S. P. Hagenaars, C. John, A. T. Williams, N. Shrine, L. Moles, K. B. Hanscombe, A. Serretti, D. J. Shepherd, and R. C. Free, “Genetic and clinical characteristics of treatment-resistant depression using primary care records in two UK cohorts,” Molecular Psychiatry, vol. 26, no. 7, pp. 3363–3373, 2021.

[20] G. Fanelli, F. Benedetti, S. Kasper, J. Zohar, D. Souery, S. Montgomery, D. Albani, G. Forloni, P. Ferentinos, D. Rujescu, J. Mendlewicz, A. Serretti, and C. Fabbri, “Higher polygenic risk scores for schizophrenia may be suggestive of treatment non-response in major depressive disorder,” Progress in Neuro-Psychopharmacology and Biological Psychiatry, vol. 108, p. 110170, 2021/06/08/ 2021, doi: 10.1016/j.pnpbp.2020.110170.

[21] A. T. Amare, K. O. Schubert, F. Tekola-Ayele, Y. H. Hsu, K. Sangkuhl, G. Jenkins, R. M. Whaley, P. Barman, A. Batzler, R. B. Altman, V. Arolt, J. Brockmoller, C. H. Chen, K. Domschke, D. K. Hall-Flavin, C. J. Hong, A. Illi, Y. Ji, O. Kampman, T. Kinoshita, E. Leinonen, Y. J. Liou, T. Mushiroda, S. Nonen, M. K. Skime, L. Wang, M. Kato, Y. L. Liu, V. Praphanphoj, J. C. Stingl, W. V. Bobo, S. J. Tsai, M. Kubo, T. E. Klein, R. M. Weinshilboum, J. M. Biernacka, and B. T. Baune, “Association of the Polygenic Scores for Personality Traits and Response to Selective Serotonin Reuptake Inhibitors in Patients with Major Depressive Disorder,” Front Psychiatry, vol. 9, p. 65, 2018, doi: 10.3389/fpsyt.2018.00065.

[22] A. T. Amare, K. O. Schubert, F. Tekola-Ayele, Y.-H. Hsu, K. Sangkuhl, G. Jenkins, R. M. Whaley, P. Barman, A. Batzler, and R. B. Altman, “The association of obesity and coronary artery disease genes with response to SSRIs treatment in major depression,” Journal of Neural Transmission, vol. 126, pp. 35–45, 2019.

[23] V. S. Marshe, M. Maciukiewicz, A.-C. Hauschild, F. Islam, L. Qin, A. K. Tiwari, E. Sibille, D. M. Blumberger, J. F. Karp, and A. J. Flint, “Genome-wide analysis suggests the importance of vascular processes and neuroinflammation in late-life antidepressant response,” Translational psychiatry, vol. 11, no. 1, p. 127, 2021.

[24] K. Perlman, D. Benrimoh, S. Israel, C. Rollins, E. Brown, J. F. Tunteng, R. You, E. You, M. Tanguay-Sela, E. Snook, M. Miresco, and M. T. Berlim, “A systematic meta-review of predictors of antidepressant treatment outcome in major depressive disorder,” (in eng), J Affect Disord, vol. 243, pp. 503–515, Jan 15 2019, doi: 10.1016/j.jad.2018.09.067.

[25] N. Dusi, S. Barlati, A. Vita, and P. Brambilla, “Brain structural effects of antidepressant treatment in major depression,” Current neuropharmacology, vol. 13, no. 4, pp. 458–465, 2015.

[26] K. F. Chi, M. Korgaonkar, and S. M. Grieve, “Imaging predictors of remission to anti-depressant medications in major depressive disorder,” Journal of Affective Disorders, vol. 186, pp. 134–144, 2015/11/01/ 2015, doi: 10.1016/j.jad.2015.07.002.

[27] P. Fettes, L. Schulze, and J. Downar, “Cortico-striatal-thalamic loop circuits of the orbitofrontal cortex: promising therapeutic targets in psychiatric illness,” Frontiers in systems neuroscience, vol. 11, p. 25, 2017.

[28] S. Kuburi, A.-M. Di Passa, V. K. Tassone, R. Mahmood, A. Lalovic, K. S. Ladha, K. Dunlop, S. Rizvi, I. Demchenko, and V. Bhat, “Neuroimaging Correlates of Treatment Response with Psychedelics in Major Depressive Disorder: A Systematic Review,” Chronic Stress, vol. 6, p. 24705470221115342, 2022.

[29] M. E. Rentería, N. K. Hansell, L. T. Strike, K. L. McMahon, G. I. de Zubicaray, I. B. Hickie, P. M. Thompson, N. G. Martin, S. E. Medland, and M. J. Wright, “Genetic architecture of subcortical brain regions: common and region-specific genetic contributions,” Genes, Brain and Behavior, vol. 13, no. 8, pp. 821–830, 2014.

[30] C. L. Satizabal, H. H. Adams, D. P. Hibar, C. C. White, M. J. Knol, J. L. Stein, M. Scholz, M. Sargurupremraj, N. Jahanshad, and G. V. Roshchupkin, “Genetic architecture of subcortical brain structures in 38,851 individuals,” Nature genetics, vol. 51, no. 11, pp. 1624–1636, 2019.

[31] D. P. Hibar, H. H. Adams, N. Jahanshad, G. Chauhan, J. L. Stein, E. Hofer, M. E. Renteria, J. C. Bis, A. Arias-Vasquez, and M. K. Ikram, “Novel genetic loci associated with hippocampal volume,” Nature communications, vol. 8, no. 1, p. 13624, 2017.

[32] M. E. Thase and A. J. Rush, “When at first you don’t succeed: sequential strategies for antidepressant nonresponders,” Journal of Clinical Psychiatry, vol. 58, no. 13, pp. 23–29, 1997.

[33] A. Minelli, C. Magri, A. Barbon, C. Bonvicini, M. Segala, C. Congiu, S. Bignotti, E. Milanesi, L. Trabucchi, and N. Cattane, “Proteasome system dysregulation and treatment resistance mechanisms in major depressive disorder,” Translational psychiatry, vol. 5, no. 12, pp. e687–e687, 2015.

[34] M. Dold, L. Bartova, G. Fugger, M. M. Mitschek, C. Fabbri, A. Serretti, J. Mendlewicz, D. Souery, J. Zohar, S. Montgomery, and S. Kasper, “Pregabalin augmentation of antidepressants in major depression - results from a European multicenter study,” (in eng), J Affect Disord, vol. 296, pp. 485–492, Jan 1 2022, doi: 10.1016/j.jad.2021.09.063.

[35] B. T. Baune, U. Dannlowski, K. Domschke, D. G. Janssen, M. A. Jordan, P. Ohrmann, J. Bauer, E. Biros, V. Arolt, and H. Kugel, “The interleukin 1 beta (IL1B) gene is associated with failure to achieve remission and impaired emotion processing in major depression,” Biological psychiatry, vol. 67, no. 6, pp. 543–549, 2010.

[36] R. H. Howland, “Sequenced treatment alternatives to relieve depression (STAR* D)--Part 2: Study outcomes,” Journal of psychosocial nursing and mental health services, vol. 46, no. 10, pp. 21–24, 2008.

[37] A. Tammiste, T. Jiang, K. Fischer, R. Mägi, K. Krjutškov, K. Pettai, T. Esko, Y. Li, K. E. Tansey, and L. S. Carroll, “Whole-exome sequencing identifies a polymorphism in the BMP5 gene associated with SSRI treatment response in major depression,” Journal of Psychopharmacology, vol. 27, no. 10, pp. 915–920, 2013.

[38] M. Lam, S. Awasthi, H. J. Watson, J. Goldstein, G. Panagiotaropoulou, V. Trubetskoy, R. Karlsson, O. Frei, C.-C. Fan, and W. De Witte, “RICOPILI: rapid imputation for COnsortias PIpeLIne,” Bioinformatics, vol. 36, no. 3, pp. 930–933, 2020.

[39] S. Das, L. Forer, S. Schönherr, C. Sidore, A. E. Locke, A. Kwong, S. I. Vrieze, E. Y. Chew, S. Levy, and M. McGue, “Next-generation genotype imputation service and methods,” Nature genetics, vol. 48, no. 10, pp. 1284–1287, 2016.

[40] T. Ge, C. Y. Chen, Y. Ni, Y. A. Feng, and J. W. Smoller, “Polygenic prediction via Bayesian regression and continuous shrinkage priors,” (in eng), Nat Commun, vol. 10, no. 1, p. 1776, Apr 16 2019, doi: 10.1038/s41467-019-09718-5.

[41] W. Viechtbauer, “Conducting meta-analyses in R with the metafor package,” Journal of statistical software, vol. 36, no. 3, pp. 1–48, 2010.

[42] D. A. Harville, “Maximum likelihood approaches to variance component estimation and to related problems,” Journal of the American statistical association, vol. 72, no. 358, pp. 320–338, 1977.

[43] W. G. Cochran, “The comparison of percentages in matched samples,” Biometrika, vol. 37, no. 3/4, pp. 256–266, 1950.

[44] J. P. Higgins, J. Thomas, J. Chandler, M. Cumpston, T. Li, M. J. Page, and V. A. Welch, Cochrane handbook for systematic reviews of interventions. John Wiley & Sons, 2019.

[45] W. Viechtbauer and M. W. L. Cheung, “Outlier and influence diagnostics for meta-analysis,” Research synthesis methods, vol. 1, no. 2, pp. 112–125, 2010.

[46] L. Palla and F. Dudbridge, “A fast method that uses polygenic scores to estimate the variance explained by genome-wide marker panels and the proportion of variants affecting a trait,” The American Journal of Human Genetics, vol. 97, no. 2, pp. 250–259, 2015.

[47] I. H. Gotlib and J. Joormann, “Cognition and depression: current status and future directions,” (in eng), Annu Rev Clin Psychol, vol. 6, pp. 285–312, 2010, doi: 10.1146/annurev.clinpsy.121208.131305.

[48] E. Bora, B. J. Harrison, C. G. Davey, M. Yücel, and C. Pantelis, “Meta-analysis of volumetric abnormalities in cortico-striatal-pallidal-thalamic circuits in major depressive disorder,” (in eng), Psychol Med, vol. 42, no. 4, pp. 671–81, Apr 2012, doi: 10.1017/s0033291711001668.

[49] M. J. Kim, J. P. Hamilton, and I. H. Gotlib, “Reduced caudate gray matter volume in women with major depressive disorder,” (in eng), Psychiatry Res, vol. 164, no. 2, pp. 114–22, Nov 30 2008, doi: 10.1016/j.pscychresns.2007.12.020.

[50] A. Khundakar, C. Morris, A. Oakley, and A. J. Thomas, “Morphometric analysis of neuronal and glial cell pathology in the caudate nucleus in late-life depression,” (in eng), Am J Geriatr Psychiatry, vol. 19, no. 2, pp. 132–41, Feb 2011, doi: 10.1097/JGP.0b013e3181df4642.

[51] M. A. Butters, H. J. Aizenstein, K. M. Hayashi, C. C. Meltzer, J. Seaman, C. F. Reynolds, A. W. Toga, P. M. Thompson, and J. T. Becker, “Three-Dimensional Surface Mapping of the Caudate Nucleus in Late-Life Depression,” The American Journal of Geriatric Psychiatry, vol. 17, no. 1, pp. 4–12, 2009/01/01/ 2009, doi: 10.1097/JGP.0b013e31816ff72b.

[52] S. S. Pillay, P. F. Renshaw, C. M. Bonello, B. C. Lafer, M. Fava, and D. Yurgelun-Todd, “A quantitative magnetic resonance imaging study of caudate and lenticular nucleus gray matter volume in primary unipolar major depression: relationship to treatment response and clinical severity,” (in eng), Psychiatry Res, vol. 84, no. 2-3, pp. 61–74, Dec 14 1998, doi: 10.1016/s0925-4927(98)00048-1.

[53] P. Willner, J. Scheel-Krüger, and C. Belzung, “The neurobiology of depression and antidepressant action,” (in eng), Neurosci Biobehav Rev, vol. 37, no. 10 Pt 1, pp. 2331–71, Dec 2013, doi: 10.1016/j.neubiorev.2012.12.007.

[54] D. A. Pizzagalli, A. J. Holmes, D. G. Dillon, E. L. Goetz, J. L. Birk, R. Bogdan, D. D. Dougherty, D. V. Iosifescu, S. L. Rauch, and M. Fava, “Reduced caudate and nucleus accumbens response to rewards in unmedicated individuals with major depressive disorder,” (in eng), Am J Psychiatry, vol. 166, no. 6, pp. 702–10, Jun 2009, doi: 10.1176/appi.ajp.2008.08081201.

[55] T. Doi, Y. Fan, J. I. Gold, and L. Ding, “The caudate nucleus contributes causally to decisions that balance reward and uncertain visual information,” eLife, vol. 9, p. e56694, 2020/06/22 2020, doi: 10.7554/eLife.56694.

[56] T. D. Satterthwaite, J. W. Kable, L. Vandekar, N. Katchmar, D. S. Bassett, C. F. Baldassano, K. Ruparel, M. A. Elliott, Y. I. Sheline, and R. C. Gur, “Common and dissociable dysfunction of the reward system in bipolar and unipolar depression,” Neuropsychopharmacology, vol. 40, no. 9, pp. 2258–2268, 2015.

[57] M. Peciña, M. Sikora, E. T. Avery, J. Heffernan, S. Peciña, B. J. Mickey, and J. K. Zubieta, “Striatal dopamine D2/3 receptor-mediated neurotransmission in major depression: Implications for anhedonia, anxiety and treatment response,” (in eng), Eur Neuropsychopharmacol, vol. 27, no. 10, pp. 977–986, Oct 2017, doi: 10.1016/j.euroneuro.2017.08.427.

[58] J. Werme, E. P. Tissink, S. C. d. Lange, M. P. v. d. Heuvel, D. Posthuma, and C. A. d. Leeuw, “Local genetic correlation analysis links depression with molecular and brain imaging endophenotypes,” medRxiv, p. 2023.03.01.23286613, 2023, doi: 10.1101/2023.03.01.23286613.

[59] F. Corponi, C. Fabbri, I. Bitter, S. Montgomery, E. Vieta, S. Kasper, S. Pallanti, and A. Serretti, “Novel antipsychotics specificity profile: A clinically oriented review of lurasidone, brexpiprazole, cariprazine and lumateperone,” European Neuropsychopharmacology, vol. 29, no. 9, pp. 971–985, 2019.

[60] J. C. Jakobsen, C. Gluud, and I. Kirsch, “Should antidepressants be used for major depressive disorder?,” BMJ Evidence-Based Medicine, vol. 25, no. 4, pp. 130–130, 2020.

[61] G. Šimić, M. Tkalčić, V. Vukić, D. Mulc, E. Španić, M. Šagud, F. E. Olucha-Bordonau, M. Vukšić, and R. H. P, “Understanding Emotions: Origins and Roles of the Amygdala,” (in eng), Biomolecules, vol. 11, no. 6, May 31 2021, doi: 10.3390/biom11060823.

[62] J. P. Hamilton, M. Siemer, and I. H. Gotlib, “Amygdala volume in major depressive disorder: a meta-analysis of magnetic resonance imaging studies,” Molecular psychiatry, vol. 13, no. 11, pp. 993–1000, 2008.

[63] K. Rajeev and C. Jonathan, “Depression: an inflammatory illness?,” Journal of Neurology, Neurosurgery & Psychiatry, vol. 83, no. 5, p. 495, 2012, doi: 10.1136/jnnp-2011-301779.

[64] A. H. Kemp, E. Gordon, A. J. Rush, and L. M. Williams, “Improving the prediction of treatment response in depression: integration of clinical, cognitive, psychophysiological, neuroimaging, and genetic measures,” CNS spectrums, vol. 13, no. 12, pp. 1066–1086, 2008.

[65] G. Laje, R. H. Perlis, A. J. Rush, and F. J. McMahon, “Pharmacogenetics studies in STAR*D: strengths, limitations, and results,” (in eng), Psychiatr Serv, vol. 60, no. 11, pp. 1446–57, Nov 2009, doi: 10.1176/appi.ps.60.11.1446.

[66] K. H. Taber, C. Wen, A. Khan, and R. A. Hurley, “The limbic thalamus,” The Journal of neuropsychiatry and clinical neurosciences, vol. 16, no. 2, pp. 127–132, 2004.

[67] M. Y. Du, Q. Z. Wu, Q. Yue, J. Li, Y. Liao, W. H. Kuang, X. Q. Huang, R. C. Chan, A. Mechelli, and Q. Y. Gong, “Voxelwise meta-analysis of gray matter reduction in major depressive disorder,” (in eng), Prog Neuropsychopharmacol Biol Psychiatry, vol. 36, no. 1, pp. 11–6, Jan 10 2012, doi: 10.1016/j.pnpbp.2011.09.014.

[68] W. Peng, Z. Chen, L. Yin, Z. Jia, and Q. Gong, “Essential brain structural alterations in major depressive disorder: a voxel-wise meta-analysis on first episode, medication-naive patients,” Journal of affective disorders, vol. 199, pp. 114–123, 2016.

[69] L. Sforzini, C. Worrell, M. Kose, I. M. Anderson, B. Aouizerate, V. Arolt, M. Bauer, B. T. Baune, P. Blier, A. J. Cleare, P. J. Cowen, T. G. Dinan, A. Fagiolini, I. N. Ferrier, U. Hegerl, A. D. Krystal, M. Leboyer, R. H. McAllister-Williams, R. S. McIntyre, A. Meyer-Lindenberg, A. H. Miller, C. B. Nemeroff, C. Normann, D. Nutt, S. Pallanti, L. Pani, B. Penninx, A. F. Schatzberg, R. C. Shelton, L. N. Yatham, A. H. Young, R. Zahn, G. Aislaitner, F. Butlen-Ducuing, C. Fletcher, M. Haberkamp, T. Laughren, F. L. Mäntylä, K. Schruers, A. Thomson, G. Arteaga-Henríquez, F. Benedetti, L. Cash-Gibson, W. R. Chae, H. De Smedt, S. M. Gold, W. J. G. Hoogendijk, V. J. Mondragón, E. Maron, J. Martynowicz, E. Melloni, C. Otte, G. Perez-Fuentes, S. Poletti, M. E. Schmidt, E. van de Ketterij, K. Woo, Y. Flossbach, J. A. Ramos-Quiroga, A. J. Savitz, and C. M. Pariante, “A Delphi-method-based consensus guideline for definition of treatment-resistant depression for clinical trials,” (in eng), Mol Psychiatry, vol. 27, no. 3, pp. 1286–1299, Mar 2022, doi: 10.1038/s41380-021-01381-x.

