## Supplementary materials for "Polygenic scores of subcortical brain volumes as possible modulators of treatment response in depression"

### Abbreviations used in Supplementary Materials

Beta, estimated coefficient of the model;

CI, Confidence intervals;

GSRD, European Group for the Study of Resistant Depression;

$I^2$, Higgin and Thompson's $I^2$ estimating how much of the total variability in the effect size estimates can be attributed to heterogeneity among the true effects;

OR, Odds ratio;

PGS, polygenic score;

pseudo-$R^2$, Nagelkerke's $R^2$;

Qp, p-value for the Cochran's Q-test of (residual) heterogeneity;

SE, standard error;

SNP, single nucleotide polymorphism;

STAR*D, Sequenced Treatment Alternatives to Relieve Depression.

### Power Analysis

Statistical power was estimated using the AVENGEME R-package (Dudbridge, 2013; Palla and Dudbridge, 2015).

SNP-based heritability ($h^2_{SNP}$) for the base traits was taken from the respective papers used to estimate PGSs weights (Satizabal *et al.*, 2019) (Hibar *et al.*, 2017). $h^2_{SNP}$ for the target traits was derived from a previous study (Pain *et al.*, 2022). The genetic covariance (cov12) between the base and target phenotypes was hypothesised to be 25% or 50%, in accordance with previous studies (Fanelli *et al.*, 2021).

The results of the power analyses are reported in the following table.

| | if cov12=0.25 | if cov12=0.5 | if cov12=0.25 | if cov12=0.5 |
|---|---|---|---|---|
| **Base trait** | **power non-response** | **power non-response** | **power non-remission** | **power non-remission** |
| Nucleus accumbens | 0.4 | 0.99 | 0.31 | 0.96 |
| Amygdala | 0.44 | 0.99 | 0.34 | 0.97 |
| Caudate nucleus | 0.48 | 1 | 0.37 | 0.98 |
| Hippocampus | 0.31 | 0.96 | 0.23 | 0.9 |
| Globus pallidus | 0.44 | 0.99 | 0.34 | 0.97 |
| Putamen | 0.48 | 1 | 0.37 | 0.98 |
| Thalamus | 0.44 | 0.99 | 0.34 | 0.97 |

### Logistic regression results

#### 1. Non-response

**Brescia sample 1**

| PGS | OR | 95% CI | Beta | SE | p-value | pseudo $R^2$ |
|---|---|---|---|---|---|---|
| Nucleus accumbens | 1.415 | 0.862, 2.323 | 0.347 | 0.253 | 0.17 | 0.022 |
| Amygdala | 1.342 | 0.825, 2.182 | 0.294 | 0.248 | 0.236 | 0.016 |
| Caudate nucleus | 1.185 | 0.738, 1.905 | 0.17 | 0.242 | 0.481 | 0.005 |
| Hippocampus | 0.93 | 0.566, 1.526 | -0.073 | 0.253 | 0.774 | 9.05e-04 |
| **Globus pallidus** | **1.808** | **1.073, 3.045** | **0.592** | **0.266** | **0.026** | **0.057** |
| Putamen | 1.182 | 0.727, 1.921 | 0.167 | 0.248 | 0.501 | 0.005 |
| Thalamus | 1.57 | 0.964, 2.558 | 0.451 | 0.249 | 0.071 | 0.036 |

**Brescia sample 2**

| PGS | OR | 95% CI | Beta | SE | p-value | pseudo $R^2$ |
|---|---|---|---|---|---|---|
| Nucleus accumbens | 0.839 | 0.602, 1.168 | -0.176 | 0.169 | 0.299 | 0.006 |
| Amygdala | 0.992 | 0.708, 1.39 | -0.008 | 0.172 | 0.963 | 1.14e-05 |
| **Caudate nucleus** | **0.683** | **0.481, 0.97** | **-0.381** | **0.179** | **0.033** | **0.024** |
| Hippocampus | 0.98 | 0.693, 1.387 | -0.02 | 0.177 | 0.908 | 6.95e-05 |
| Globus pallidus | 1.12 | 0.798, 1.572 | 0.113 | 0.173 | 0.513 | 0.002 |
| Putamen | 0.867 | 0.616, 1.219 | -0.143 | 0.174 | 0.41 | 0.004 |
| Thalamus | 0.848 | 0.598, 1.202 | -0.165 | 0.178 | 0.355 | 0.005 |

**GSRD**

| PGS | OR | 95% CI | Beta | SE | p-value | pseudo $R^2$ |
|---|---|---|---|---|---|---|
| Nucleus accumbens | 1.019 | 0.883, 1.176 | 0.019 | 0.073 | 0.795 | 8.08e-05 |
| Amygdala | 1.119 | 0.969, 1.291 | 0.112 | 0.073 | 0.126 | 0.003 |
| Caudate nucleus | 1.093 | 0.946, 1.264 | 0.089 | 0.074 | 0.225 | 0.002 |
| Hippocampus | 1.067 | 0.916, 1.243 | 0.065 | 0.078 | 0.406 | 0.001 |
| Globus pallidus | 1.022 | 0.884, 1.182 | 0.022 | 0.074 | 0.77 | 1.03e-04 |
| Putamen | 0.947 | 0.808, 1.11 | -0.054 | 0.081 | 0.504 | 0.001 |
| Thalamus | 0.918 | 0.795, 1.059 | -0.086 | 0.073 | 0.235 | 0.002 |

**Münster**

| PGS | OR | 95% CI | Beta | SE | p-value | pseudo $R^2$ |
|---|---|---|---|---|---|---|
| Nucleus accumbens | 0.988 | 0.823, 1.186 | -0.012 | 0.093 | 0.898 | 3.86e-05 |
| Amygdala | 0.921 | 0.771, 1.101 | -0.082 | 0.091 | 0.366 | 0.002 |
| Caudate nucleus | 0.939 | 0.782, 1.127 | -0.063 | 0.093 | 0.499 | 0.001 |
| Hippocampus | 1.116 | 0.932, 1.337 | 0.11 | 0.092 | 0.233 | 0.003 |
| Globus pallidus | 0.85 | 0.709, 1.017 | -0.163 | 0.092 | 0.076 | 0.007 |
| Putamen | 0.937 | 0.779, 1.127 | -0.065 | 0.094 | 0.49 | 0.001 |
| **Thalamus** | **0.832** | **0.695, 0.996** | **-0.184** | **0.092** | **0.044** | **0.01** |

**STAR*D**

| PGS | OR | 95% CI | Beta | SE | p-value | pseudo $R^2$ |
|---|---|---|---|---|---|---|
| Nucleus accumbens | 0.974 | 0.871, 1.09 | -0.026 | 0.057 | 0.649 | 1.88e-04 |
| Amygdala | 1.065 | 0.949, 1.196 | 0.063 | 0.059 | 0.282 | 0.001 |
| Caudate nucleus | 1.042 | 0.93, 1.167 | 0.041 | 0.058 | 0.484 | 4.45e-04 |
| Hippocampus | 0.987 | 0.881, 1.106 | -0.013 | 0.058 | 0.817 | 4.88e-05 |
| Globus pallidus | 1.078 | 0.964, 1.205 | 0.075 | 0.057 | 0.186 | 0.002 |
| Putamen | 1.055 | 0.935, 1.192 | 0.054 | 0.062 | 0.388 | 0.002 |
| Thalamus | 0.987 | 0.881, 1.106 | -0.013 | 0.058 | 0.82 | 4.70e-05 |

**Tartu**

| PGS | OR | 95% CI | Beta | SE | p-value | pseudo $R^2$ |
|---|---|---|---|---|---|---|
| Nucleus accumbens | 1.537 | 0.786, 3.005 | 0.43 | 0.342 | 0.208 | 0.023 |
| Amygdala | 1.255 | 0.662, 2.377 | 0.227 | 0.326 | 0.486 | 0.007 |
| Caudate nucleus | 0.98 | 0.505, 1.901 | -0.02 | 0.338 | 0.953 | 4.68e-05 |
| Hippocampus | 0.665 | 0.348, 1.27 | -0.408 | 0.33 | 0.216 | 0.022 |
| Globus pallidus | 1.613 | 0.8, 3.253 | 0.478 | 0.358 | 0.182 | 0.025 |
| Putamen | 1.401 | 0.753, 2.607 | 0.337 | 0.317 | 0.287 | 0.016 |
| Thalamus | 0.893 | 0.438, 1.819 | -0.113 | 0.363 | 0.756 | 0.001 |

Results with p-value <0.05 are in bold.

### 2. Non remission

#### GSRD

| PGS | OR | 95% CI | Beta | SE | p-value | pseudo $R^2$ |
|---|---|---|---|---|---|---|
| Nucleus accumbens | 1.08 | 0.913, 1.278 | 0.077 | 0.086 | 0.371 | 0.001 |
| Amygdala | 1.081 | 0.913, 1.28 | 0.078 | 0.086 | 0.365 | 0.001 |
| **Caudate nucleus** | **1.202** | **1.012, 1.428** | **0.184** | **0.088** | **0.036** | **0.006** |
| Hippocampus | 1.024 | 0.859, 1.222 | 0.024 | 0.09 | 0.793 | 9.18e-05 |
| Globus pallidus | 1.09 | 0.921, 1.29 | 0.086 | 0.086 | 0.319 | 0.001 |
| Putamen | 0.997 | 0.828, 1.201 | -0.003 | 0.095 | 0.979 | 9.64e-07 |
| Thalamus | 0.897 | 0.759, 1.059 | -0.109 | 0.085 | 0.198 | 0.002 |

#### Münster

| PGS | OR | 95% CI | Beta | SE | p-value | pseudo $R^2$ |
|---|---|---|---|---|---|---|
| Nucleus accumbens | 0.969 | 0.81, 1.161 | -0.031 | 0.092 | 0.738 | 2.52e-04 |
| Amygdala | 0.916 | 0.768, 1.092 | -0.088 | 0.09 | 0.327 | 0.002 |
| Caudate nucleus | 0.98 | 0.82, 1.172 | -0.02 | 0.091 | 0.826 | 1.08e-04 |
| Hippocampus | 0.966 | 0.808, 1.154 | -0.035 | 0.091 | 0.703 | 3.27e-04 |
| Globus pallidus | 0.858 | 0.718, 1.026 | -0.153 | 0.091 | 0.095 | 0.006 |
| Putamen | 0.918 | 0.765, 1.101 | -0.086 | 0.093 | 0.357 | 0.002 |
| **Thalamus** | **0.797** | **0.668, 0.951** | **-0.227** | **0.09** | **0.012** | **0.014** |

#### STAR*D

| PGS | OR | 95% CI | Beta | SE | p-value | pseudo $R^2$ |
|---|---|---|---|---|---|---|
| Nucleus accumbens | 1.02 | 0.911, 1.143 | 0.02 | 0.058 | 0.73 | 1.04e-04 |
| Amygdala | 1.108 | 0.986, 1.247 | 0.103 | 0.06 | 0.088 | 0.003 |
| Caudate nucleus | 1.099 | 0.977, 1.236 | 0.094 | 0.06 | 0.116 | 0.002 |
| Hippocampus | 1.022 | 0.909, 1.15 | 0.022 | 0.06 | 0.708 | 1.23e-04 |
| Globus pallidus | 1.062 | 0.946, 1.192 | 0.06 | 0.059 | 0.303 | 0.001 |
| Putamen | 1.033 | 0.911, 1.171 | 0.032 | 0.064 | 0.614 | 2.26e-04 |
| Thalamus | 1.007 | 0.897, 1.13 | 0.007 | 0.059 | 0.908 | 1.16e-05 |

**Tartu**

| PGS | OR | 95% CI | Beta | SE | p-value | pseudo $R^2$ |
|---|---|---|---|---|---|---|
| Nucleus accumbens | 1.571 | 0.776, 3.182 | 0.452 | 0.36 | 0.21 | 0.021 |
| Amygdala | 1.451 | 0.721, 2.92 | 0.372 | 0.357 | 0.296 | 0.014 |
| Caudate nucleus | 1.107 | 0.553, 2.216 | 0.102 | 0.354 | 0.774 | 0.001 |
| Hippocampus | 0.803 | 0.427, 1.51 | -0.219 | 0.322 | 0.496 | 0.006 |
| Globus pallidus | 1.87 | 0.897, 3.9 | 0.626 | 0.375 | 0.095 | 0.037 |
| Putamen | 1.525 | 0.771, 3.016 | 0.422 | 0.348 | 0.225 | 0.019 |
| Thalamus | 1.025 | 0.517, 2.032 | 0.025 | 0.349 | 0.943 | 6.21e-05 |

Results with p-values <0.05 are depicted in bold.

### Leave-one-out sensitivity analyses

#### 1. Non-response

**Nucleus accumbens**

| Sample excluded | OR | 95% CI | Beta | SE | p-value | Qp | $I^2$ |
|---|---|---|---|---|---|---|---|
| Brescia sample 1 | 0.987 | 0.915, 1.066 | -0.013 | 0.039 | 0.746 | 0.583 | 0.046 |
| Brescia sample 2 | 1.005 | 0.93, 1.086 | 0.005 | 0.04 | 0.897 | 0.443 | 0.017 |
| Tartu | 0.99 | 0.918, 1.068 | -0.01 | 0.039 | 0.799 | 0.527 | 0.064 |
| GSRD | 0.987 | 0.903, 1.079 | -0.013 | 0.045 | 0.77 | 0.322 | 0.041 |
| Münster | 0.997 | 0.918, 1.084 | -0.003 | 0.042 | 0.949 | 0.307 | 0.003 |
| STAR*D | 1.014 | 0.915, 1.124 | 0.014 | 0.053 | 0.787 | 0.336 | 0.016 |

**Amygdala**

| Sample excluded | OR | 95% CI | Beta | SE | p-value | Qp | $I^2$ |
|---|---|---|---|---|---|---|---|
| Brescia sample 1 | 1.05 | 0.971, 1.134 | 0.049 | 0.04 | 0.22 | 0.51 | 0 |
| Brescia sample 2 | 1.06 | 0.979, 1.147 | 0.058 | 0.04 | 0.149 | 0.391 | 0.352 |
| Tartu | 1.053 | 0.975, 1.138 | 0.052 | 0.039 | 0.185 | 0.411 | 0 |
| GSRD | 1.031 | 0.934, 1.138 | 0.031 | 0.05 | 0.543 | 0.495 | 5.474 |
| **Münster** | **1.089** | **1.001, 1.186** | **0.086** | **0.043** | **0.048** | **0.832** | **0** |
| STAR*D | 1.049 | 0.921, 1.196 | 0.048 | 0.067 | 0.47 | 0.378 | 22.341 |

**Caudate nucleus**

| Sample excluded | OR | 95% CI | Beta | SE | p-value | Qp | $I^2$ |
|---|---|---|---|---|---|---|---|
| Brescia sample 1 | 0.995 | 0.892, 1.11 | -0.005 | 0.056 | 0.924 | 0.145 | 34.929 |
| Brescia sample 2 | 1.039 | 0.961, 1.124 | 0.039 | 0.04 | 0.337 | 0.736 | 0 |
| Tartu | 1.01 | 0.919, 1.11 | 0.01 | 0.048 | 0.837 | 0.124 | 21.449 |
| GSRD | 0.964 | 0.839, 1.108 | -0.037 | 0.071 | 0.602 | 0.203 | 32.865 |
| Münster | 1.037 | 0.952, 1.129 | 0.036 | 0.043 | 0.404 | 0.177 | 0.015 |
| STAR*D | 0.968 | 0.815, 1.15 | -0.032 | 0.088 | 0.714 | 0.137 | 46.723 |

**Hippocampus**

| Sample excluded | OR | 95% CI | Beta | SE | p-value | Qp | $I^2$ |
|---|---|---|---|---|---|---|---|
| Brescia sample 1 | 1.025 | 0.947, 1.109 | 0.018 | 0.057 | 0.751 | 0.146 | 38.369 |
| Brescia sample 2 | 1.025 | 0.946, 1.11 | 0.07 | 0.095 | 0.462 | 0.026 | 72.996 |
| Tartu | 1.029 | 0.951, 1.113 | 0.037 | 0.067 | 0.579 | 0.045 | 54.262 |
| GSRD | 1.007 | 0.92, 1.102 | 0.114 | 0.119 | 0.338 | 0.024 | 72.385 |
| Münster | 1.002 | 0.92, 1.092 | 0.079 | 0.043 | 0.066 | 0.231 | 0.017 |
| STAR*D | 1.055 | 0.948, 1.173 | 0.1 | 0.121 | 0.41 | 0.034 | 70.234 |

**Globus pallidus**

| Sample excluded | OR | 95% CI | Beta | SE | p-value | Qp | $I^2$ |
|---|---|---|---|---|---|---|---|
| Brescia sample 1 | 1.018 | 0.91, 1.139 | 0.024 | 0.04 | 0.543 | 0.503 | 0.019 |
| Brescia sample 2 | 1.073 | 0.89, 1.293 | 0.024 | 0.041 | 0.55 | 0.49 | 0.042 |
| Tartu | 1.038 | 0.91, 1.184 | 0.028 | 0.04 | 0.478 | 0.78 | 0 |
| GSRD | 1.12 | 0.888, 1.413 | 0.007 | 0.046 | 0.876 | 0.545 | 0 |
| Münster | 1.082 | 0.995, 1.176 | 0.002 | 0.044 | 0.961 | 0.669 | 0 |
| STAR*D | 1.105 | 0.872, 1.4 | 0.053 | 0.054 | 0.327 | 0.596 | 0 |

**Putamen**

| Sample excluded | OR | 95% CI | Beta | SE | p-value | Qp | $I^2$ |
|---|---|---|---|---|---|---|---|
| Brescia sample 1 | 0.995 | 0.916, 1.08 | -0.005 | 0.042 | 0.898 | 0.484 | 0.003 |
| Brescia sample 2 | 1.008 | 0.927, 1.096 | 0.008 | 0.043 | 0.851 | 0.522 | 0 |
| Tartu | 0.994 | 0.915, 1.079 | -0.006 | 0.042 | 0.877 | 0.596 | 0 |
| GSRD | 1.019 | 0.927, 1.12 | 0.018 | 0.048 | 0.703 | 0.503 | 0 |
| Münster | 1.015 | 0.927, 1.111 | 0.015 | 0.046 | 0.747 | 0.501 | 0 |
| STAR*D | 0.957 | 0.858, 1.067 | -0.044 | 0.056 | 0.429 | 0.633 | 0 |

**Thalamus**

| Sample excluded | OR | 95% CI | Beta | SE | p-value | Qp | $I^2$ |
|---|---|---|---|---|---|---|---|
| Brescia sample 1 | 0.924 | 0.849, 1.006 | -0.079 | 0.043 | 0.068 | 0.58 | 10.037 |
| Brescia sample 2 | 0.943 | 0.861, 1.034 | -0.058 | 0.047 | 0.215 | 0.144 | 15.922 |
| Tartu | 0.938 | 0.861, 1.023 | -0.064 | 0.044 | 0.15 | 0.127 | 13.012 |
| GSRD | 0.946 | 0.82, 1.091 | -0.055 | 0.073 | 0.447 | 0.134 | 35.364 |
| Münster | 0.966 | 0.888, 1.05 | -0.035 | 0.043 | 0.415 | 0.285 | 0.03 |
| STAR*D | 0.903 | 0.815, 1.001 | -0.102 | 0.052 | 0.052 | 0.207 | 0.024 |

Results that are different from the main analysis are reported in bold (i.e., that changed from being nominally significant to be non-significant or vice versa).

2. **Non-remission**

**Nucleus accumbens**

| Sample excluded | OR | 95% CI | Beta | SE | p-value | Qp | $I^2$ |
|---|---|---|---|---|---|---|---|
| Tartu | 1.023 | 0.942, 1.112 | 0.023 | 0.042 | 0.588 | 0.691 | 0 |
| GSRD | 1.014 | 0.922, 1.115 | 0.014 | 0.049 | 0.779 | 0.422 | 0.019 |
| Münster | 1.046 | 0.953, 1.148 | 0.045 | 0.048 | 0.344 | 0.45 | 0.034 |
| STAR*D | 1.04 | 0.921, 1.173 | 0.039 | 0.062 | 0.527 | 0.353 | 0.022 |

**Amygdala**

| Sample excluded | OR | 95% CI | Beta | SE | p-value | Qp | $I^2$ |
|---|---|---|---|---|---|---|---|
| Tartu | 1.045 | 0.936, 1.166 | 0.044 | 0.056 | 0.438 | 0.198 | 37.134 |
| GSRD | 1.041 | 0.876, 2.237 | 0.04 | 0.088 | 0.651 | 0.139 | 50.028 |
| **Münster** | **1.105** | **1.004, 1.216** | **0.1** | **0.049** | **0.041** | **0.721** | **0** |
| STAR*D | 1.015 | 0.868, 1.188 | 0.015 | 0.08 | 0.85 | 0.241 | 29.316 |

**Caudate nucleus**

| Sample excluded | OR | 95% CI | Beta | SE | p-value | Qp | $I^2$ |
|---|---|---|---|---|---|---|---|
| Tartu | 1.094 | 1.003, 1.194 | 0.09 | 0.044 | 0.043 | 0.27 | 3.29 |
| **GSRD** | **1.062** | **0.963, 1.171** | **0.06** | **0.05** | **0.23** | **0.572** | **0.779** |
| Münster | 1.13 | 1.027, 1.245 | 0.123 | 0.049 | 0.012 | 0.7 | 0 |
| **STAR*D** | **1.088** | **0.911, 1.3** | **0.085** | **0.091** | **0.351** | **0.271** | **39.71** |

**Hippocampus**

| Sample excluded | OR | 95% CI | Beta | SE | p-value | Qp | $I^2$ |
|---|---|---|---|---|---|---|---|
| Tartu | 1.01 | 0.927, 1.1 | 0.007 | 0.069 | 0.922 | 0.099 | 58.023 |
| GSRD | 1 | 0.908, 1.101 | 0.021 | 0.123 | 0.863 | 0.037 | 71.282 |
| Münster | 1.017 | 0.924, 1.12 | 0.077 | 0.048 | 0.107 | 0.328 | 0.018 |
| STAR*D | 0.987 | 0.872, 1.116 | 0.043 | 0.143 | 0.765 | 0.038 | 72.43 |

**Globus pallidus**

| Sample excluded | OR | 95% CI | Beta | SE | p-value | Qp | $I^2$ |
|---|---|---|---|---|---|---|---|
| Tartu | 1.007 | 0.879, 2.153 | 0.01 | 0.044 | 0.827 | 0.858 | 0 |
| GSRD | 1.021 | 0.803, 3.299 | 0 | 0.049 | 1 | 0.688 | 0 |
| Münster | 1.08 | 0.984, 1.187 | 0.017 | 0.049 | 0.728 | 0.759 | 0 |
| STAR*D | 1.044 | 0.789, 3.381 | -0.013 | 0.063 | 0.83 | 0.729 | 0 |

**Putamen**

| Sample excluded | OR | 95% CI | Beta | SE | p-value | Qp | $I^2$ |
|---|---|---|---|---|---|---|---|
| Tartu | 0.995 | 0.909, 1.089 | -0.005 | 0.046 | 0.917 | 0.579 | 0 |
| GSRD | 1.004 | 0.907, 1.112 | 0.004 | 0.052 | 0.937 | 0.277 | 0.052 |
| Münster | 1.031 | 0.93, 1.142 | 0.03 | 0.052 | 0.561 | 0.5 | 0.013 |
| STAR*D | 0.972 | 0.855, 1.105 | -0.029 | 0.065 | 0.663 | 0.345 | 0.024 |

**Thalamus**

| Sample excluded | OR | 95% CI | Beta | SE | p-value | Qp | $I^2$ |
|---|---|---|---|---|---|---|---|
| Tartu | 0.908 | 0.792, 2.04 | -0.097 | 0.07 | 0.164 | 0.086 | 58.613 |
| GSRD | 0.915 | 0.75, 2.116 | -0.089 | 0.101 | 0.38 | 0.093 | 60.31 |
| Münster | 0.968 | 0.873, 1.073 | -0.033 | 0.053 | 0.535 | 0.526 | 7.581 |
| **STAR*D** | **0.853** | **0.757, 0.961** | **-0.159** | **0.061** | **0.009** | **0.552** | **0** |

Results that are different from the main analysis are reported in bold (i.e., that changed from being nominally significant to be non-significant or vice versa).

### References

Dudbridge F. (2013). Power and predictive accuracy of polygenic risk scores. *PLoS genetics* **9**:e1003348.

Fanelli G, Benedetti F, Kasper S, Zohar J, Souery D, Montgomery S *et al.* (2021). Higher polygenic risk scores for schizophrenia may be suggestive of treatment non-response in major depressive disorder. *Progress in Neuro-Psychopharmacology and Biological Psychiatry* **108**:110170.

Hibar DP, Adams HH, Jahanshad N, Chauhan G, Stein JL, Hofer E *et al.* (2017). Novel genetic loci associated with hippocampal volume. *Nature communications* **8**:13624.

Pain O, Hodgson K, Trubetskoy V, Ripke S, Marshe VS, Adams MJ *et al.* (2022). Identifying the common genetic basis of antidepressant response. *Biological psychiatry global open science* **2**:115-126.

Palla L, Dudbridge F. (2015). A Fast Method that Uses Polygenic Scores to Estimate the Variance Explained by Genome-wide Marker Panels and the Proportion of Variants Affecting a Trait. *The American Journal of Human Genetics* **97**:250-259.

Satizabal CL, Adams HH, Hibar DP, White CC, Knol MJ, Stein JL *et al.* (2019). Genetic architecture of subcortical brain structures in 38,851 individuals. *Nature genetics* **51**:1624-1636.
